# Genetic Testing in patients with Dementia: A Data-Driven Clinical Decision Tree for Memory Clinics

**DOI:** 10.1101/2024.01.14.24301065

**Authors:** Sven J. van der Lee, Marc Hulsman, Rosalina M.L. van Spaendonk, Jetske van der Schaar, Janna I.R. Dijkstra, Niccoló Tesi, Fred van Ruissen, Mariet W. Elting, Marcel T. Reinders, Itziar de Rojas, Corien Verschuuren, Wiesje M. van der Flier, Mieke M. van Haelst, Christa M. de Geus, Yolande A.L. Pijnenburg, Henne H. Holstege

## Abstract

**Importance:** Identifying genetic causes for dementia in patients who visit a memory clinic is important for patients and family members. However, current clinical selection criteria for genetic analysis may miss carriers of pathogenic genetic variants (PGVs) in dementia-related genes.

**Objective:** Optimizing the patient-selection criteria for offering genetic counselling in patients visiting memory clinics.

**Design:** Clinical cohort study at the Alzheimer Center Amsterdam, analysing patients from January 2010 to June 2012, and who participated in the Amsterdam Dementia Cohort. A 54-gene dementia panel was used to identify PGVs, class IV/V variants according to the American College of Medical Genetics and Genomics (ACMG) guidelines. Subsequently, we formulated a novel decision tree to determine eligibility for genetic testing, allowing optimal identification of symptomatic PGV carriers. The decision tree was prospectively applied in the same memory clinic for one year (2021-2022).

**Setting:** The Alzheimer Center Amsterdam, a specialized memory clinic in the Netherlands.

**Participants:** A total of 1,138 patients visited the memory clinic (2010-2012), of whom 1,022 were genetically analysed [90%]. Of the analysed patients 413 were female [40.4%], mean [SD] age at presentation 62.1 [8.9] years. The decision tree was applied to 517 patients that visited the memory clinic between 2021-2022; 215[41.6%] female, mean [SD] age at presentation 64.1[8.5] years.

**Exposure:** none

**Main Outcome(s):** Presence of a PGVs and eligibility of carriers for genetic testing based on previous and new clinical selection criteria.

**Results:** We identified 34 PGV carriers, corresponding to 3.3% of all patients. Of these, 24 carriers had symptoms of dementia [n=24]. Based on previous clinical criteria, only 15 of all PGV carriers were eligible. Which was 44% of all PGV carriers [15/34] and 65% of symptomatic PGV carriers [15/24]. With the new decision tree, 22 of all PGVs were eligible 62.5% [22/34] and 91% [22/24] of all symptomatic PGV carriers were eligible. In the prospective application, 517 patients were evaluated of which 148[31%] patients were eligible for a genetic test, 103 [20%] were finally tested and 13 patients carried a PGV [2.5% of total]. There were 73% more patients with a PGV identified than anticipated.

**Conclusions and Relevance:** Our decision tree improved the identification of patients with genetic dementias.

**Key Points:** *Question:* Do current clinical criteria for selecting dementia patients identify those with pathogenic genetic variants (PGVs) in dementia-related genes?

*Findings:* In a cohort study at the Alzheimer Center Amsterdam, 34 PGV carriers were identified among 1,022 patients. Previous criteria identified only 44% (15/34) of all carriers and 65% (15/24) of symptomatic carriers. A new decision tree increased this to 62.5% (22/34) and 91% (22/24), respectively. Real-life implementation improved carrier identification by 73%.

*Meaning:* Our decision tree enhances genetic dementia patient identification, offering an improved approach to identify families with familial dementia.

## Introduction

Dementia affects millions of people worldwide and places a large burden on patients, caregivers, and society as whole.^1^ While the cause of dementia is commonly a complex interplay of multiple genetic and environmental factors, a minority of patients has a monogenic (familial) form of dementia.^2^ Identifying patients with familial dementia and the associated causative pathogenic genetic variant (PGV) is difficult, yet important as it influences the diagnostic process and provides the starting point for future treatment strategies.^1,3,4^

Monogenic dementias are explained by PGVs in one of >50 dementia-related genes^5^, many of which were discovered during the last ten years, due to the advances of next-generation sequencing (NGS) techniques.^5–7^ All these PGVs have important clinical relevance. For example, PGVs in *PSEN1* can cause Alzheimer’s disease (AD)^8^ and the hexanucleotide repeat expansion in the *C9ORF*72 gene can cause frontotemporal dementia (FTD).^9^ Since these variants can explain the cause of disease in affected family members, they open up the opportunity for pre–symptomatic genetic testing of cognitively healthy relatives and preventive options in family-planning.^6^ In addition, carriership of a PGV may enable participation in clinical trials such the Dominantly Inherited Alzheimer Network Trial Unit (DIAN-TU)^10^ and the Genetic Frontotemporal dementia Initiative (GENFI).^11^ The reduced costs of NGS techniques led to the implementation of dementia gene panels to identify patients PGVs in patients with suspected genetic dementia,^12^ This has led in an increased patient-interest in knowing their genetic status.^13^

In our specialized memory clinic, 10-15% of all patients are tested and 10% of the patients carried a PGV.^2^ Patient populations visiting memory clinics are generally older, more diverse, less affected.^14,15^ Therefore, we estimate that <1% of all patients who enter a memory clinic are currently identified to have a PGV. This percentage may be higher in clinics that specialize in early-onset dementia.^16–18^ Indeed, 12.6% of a large cohort of well-defined (young onset) dementia patients and healthy aged controls carried a PGVs in one of 17 dementia-related genes^14^ . This is in stark contrast with our estimates Nevertheless, since only a minority of patients is genetically tested, often due to practical or financial related issues.^19^ it is very likely that PGV carriers in memory clinics remain unidentified. However, even if genetic testing would be broadly available, the clinical criteria applied to select patients eligible for genetic testing are (historically) strict and vary across countries and hospitals.^6,12,18,20^

The eligibility criteria for genetic testing used in clinical care involves having early onset dementia (<50 or <60 years)^6^ and/or on having a (strong) positive family history.^6,21,22^ However, not all patients carrying a PGV present with typical characteristics and often the clinical diagnosis does not match the genetic diagnosis, such that many PGV-carriers are not identified as eligible for genetic testing.^14^ Clinical criteria to determine eligibility for testing do not take this into account. For example, the National Health Service (NHS) has different clinical criteria per diagnosis of dementia.^23^ The age at onset of diseases caused by PGVs is also more variable than outlined in the criteria, and some carriers may escape disease until older ages.^14,24^ Furthermore, the absence of strong familial history does not exclude a genetic cause of dementia. Family history may be uninformative or incomplete due to small pedigree sizes, intrafamilial variability, unknown disease status of family members and premature deaths. In addition to variation in age at onset, genetic variants can have wide pleiotropy in phenotypes, masking autosomal dominant inheritance patterns and thereby concealing a family history of dementia. For example, the *C9ORF72* repeat expansion is not exclusively associated with clinically diagnosed FTD and amyotrophic lateral sclerosis (ALS)^9,25^, since carriers also present with psychiatric disorders^26^, Huntington’s disease-like syndromes^27^, and AD^28^. Lastly, the occurrence of ‘*de novo*’ mutations may completely preclude autosomal dominant inheritance patterns.^8^

In summary, despite the important implications and increasing patient-interest it is likely that many PGV carriers are not offered genetic testing and thus remain unidentified. Optimizing the patient-selection criteria for offering genetic counselling can improve detection of PGV carriers in dementia care. In this study, we attempt to improve selection criteria by first performing genetic tests for nearly *all* patients who visited a specialized memory clinic during a period of 2.5 years. We determined the prevalence of PGVs and evaluated current clinical selection criteria for genetic testing. Then, based on our results, we studied whether a simple decision tree would yield a better detection of PGVs. Lastly, we prospectively implemented this decision tree in routine clinical care and evaluated the results of the first year of implementation.

## Methods

In this paper we describe two complementary studies embedded in the Alzheimer Center Amsterdam. First, a retrospective study into the prevalence of genetic dementia in historical patient data collected between in 2010 and 2012. Findings from this study were used to formulate novel clinical selection criteria for the genetic testing, which we implemented in a prospective study in the years 2021 to 2022. The study design is summarized in **Figure 1**.

**Figure 1:**
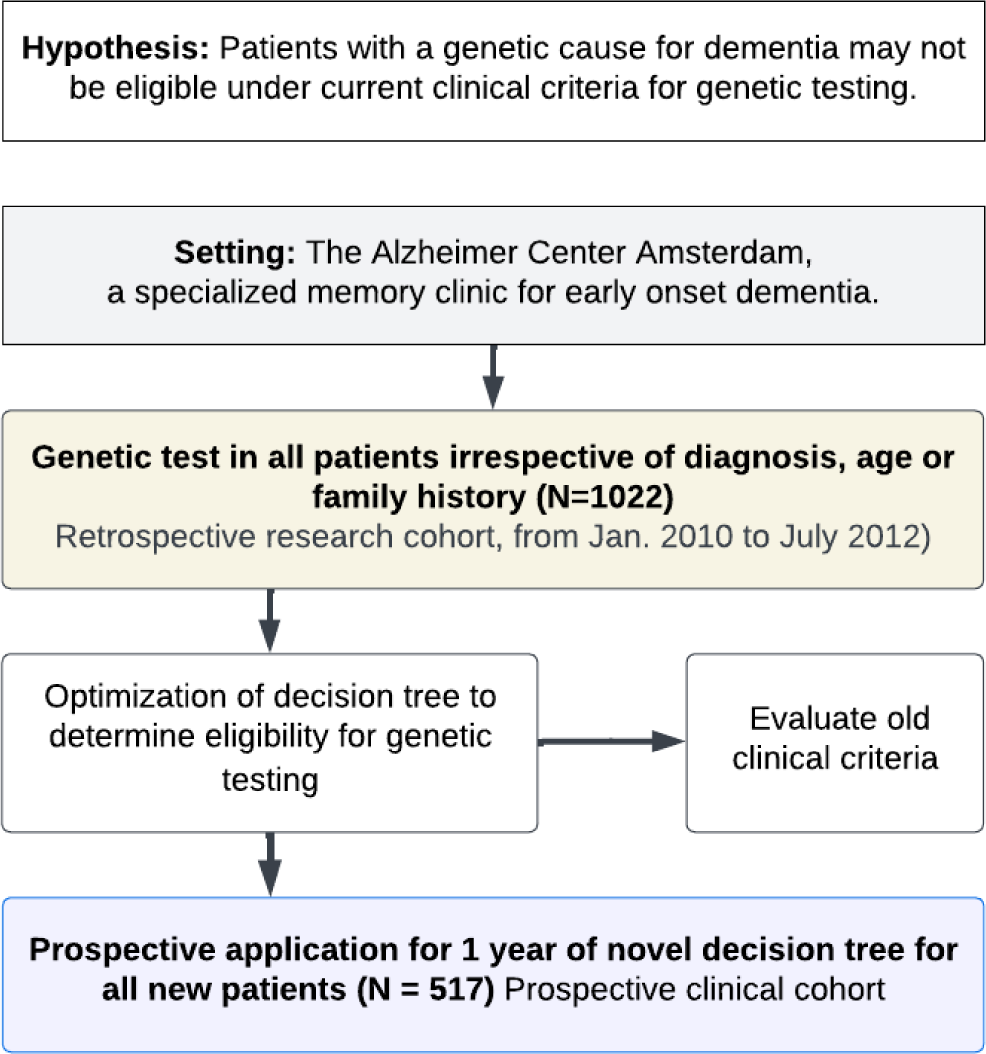
Summary of the hypothesis and studies.

### Patient population

The Alzheimer Center Amsterdam is a specialized memory clinic for young onset dementia. Patients visit the Alzheimer Center Amsterdam for an analysis of their cognitive complaints. All patients undergo the same diagnostic trajectory which did not change significantly between 2010 and 2022.^29,30^ The standardized diagnostic workup included a medical and neurological investigation, neuropsychological assessment, brain magnetic resonance imaging (MRI) and optional cerebrospinal fluid (CSF) analysis.^29,30^ The vast majority (estimated >95%) of patients consent to research in the Amsterdam Dementia Cohort (ADC).^29,30^ Patients provide informed consent for the use of their medical data for research purposes and (optional consent) for storage of their DNA in a dedicated biobank.^29^

### Study cohorts

#### Retrospective study

For this part of the study, we included 1,138 patients who consecutively visited the clinic between the January 2010 and July 2012 and who consented to use of their medical data. A genetic test was performed in 1,022 (90%) of these patients. Of the 116 not tested, 45 patients did not consent research of their DNA (4.1%), no DNA was available for 60 patients (5.5%) and 11 patients were excluded because of poor DNA or sequencing quality (1.0%). We evaluated the relevant clinical changes reported in the clinical records of all identified PGV-carriers up until the end of 2022, i.e., at least 10 years of follow-up since initial presentation. Finally, we evaluated the old eligibility-criteria (**Table 1**) for genetic testing. Notably: we applied the eligibility-criteria retrospectively, since next-generation sequencing was not implemented in the clinic in the years studied.^31^

**Table 1:** Clinical criteria to determine eligibility for genetic testing: applied from 2017 until 2021.

| Criteria list |
| --- |
| 1) AD with age at onset <60 years, |
| 2) AD with age at onset <70 years and family member with AD <70 years, |
| 3) AD phenotype without AD biomarkers |
| 4) Suspicion of (bv) FTD or PPA |
| 5) White matter lesions e.c.i. |
| 6) DLB age at onset <70 years and a family member with DLB <70 years |
| 7) Dementia with >=3 family members with dementia |

#### Prospective study

After creating a decision tree based on results from in the retrospective study, we applied it prospectively to 517 patients who visited the Alzheimer Center Amsterdam between September 2021 and September 2022 and who gave informed consent to use their medical data for research. The clinical practice was adapted as shown in **Figure 2**. The main change is that criteria for genetic testing were systematically applied in *all* patients at time of diagnosis. Four months after inclusion of the last patient in the prospective cohort, at the end of 2022, we checked all medical records and recorded 1) if counselling by a clinical geneticist had taken place, 2) if a genetic test was done, 3) if a PGV was identified or unknown significance (VUS) was reported.

**Figure 2:**
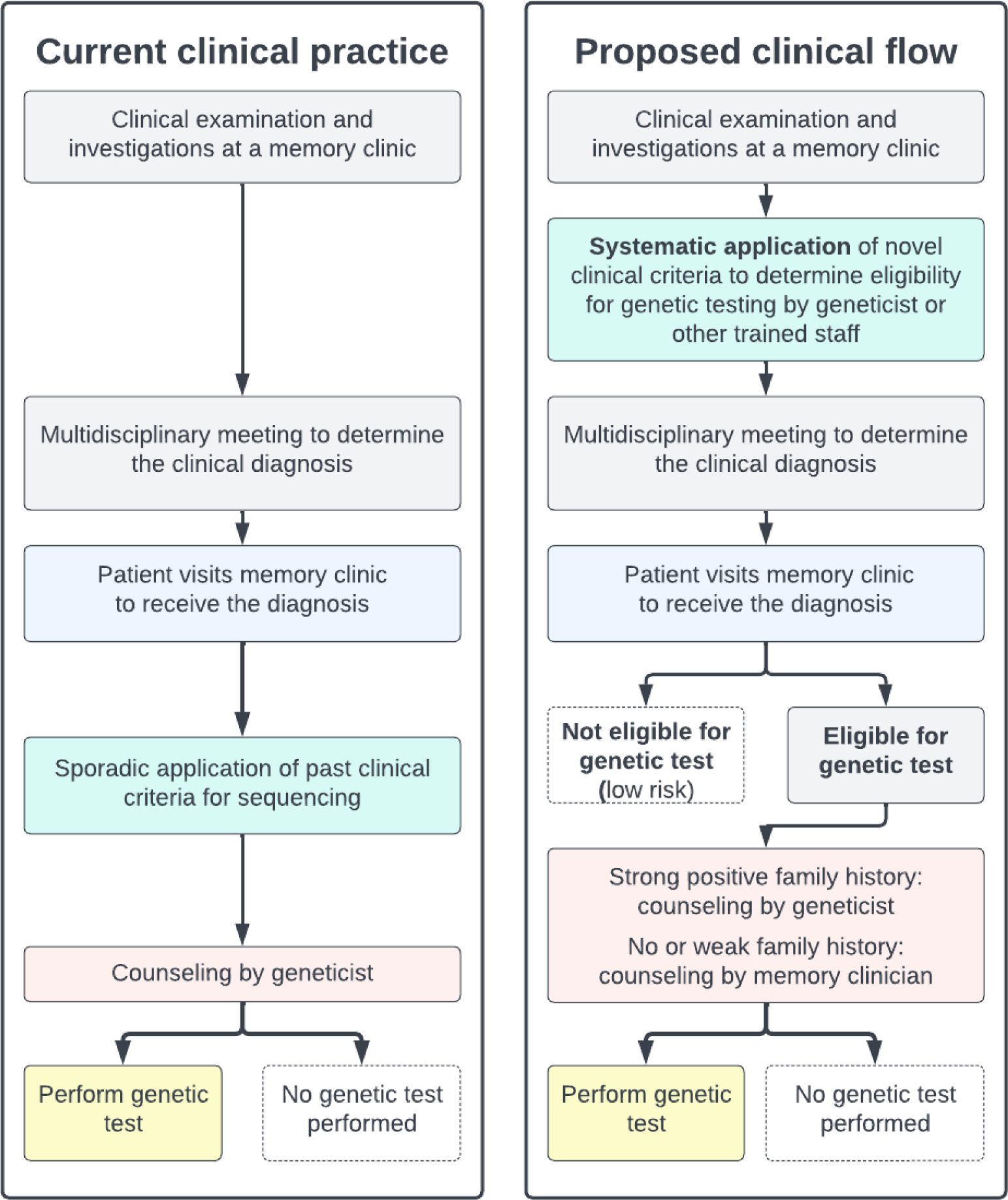
The figure shows in the left block the clinical practice in most memory clinics. Clinical criteria for genetic testing (if present) are sporadically applied to patients that visit a memory clinic. Based on current finding we propose a different clinical workflow. In this workflow the clinician asseses the eligibility for genetic testing at time of the initial diagnosis in the multidiciplinary meeting that precedes diagnosis in most memory clinics. Based on this assessment the patients are eligible, or not. If there is a strong suspicion of monogenic dementia because of clinical signs or a strong family history the patient is referred to a clinical geneticist after which testing takes place (or not). If this strong suspicion is not present the memory clinic clinician can counsel and test the patient for monogenic dementia (or not).

### Genetic tests

We tested for 1) PGVs (Class IV and V based on the ACMG)^15^ in a dementia gene panel (targeted exome-sequencing) consisting of 54-genes, 2) the *C9ORF72* hexanucleotide repeat expansion, and 3) for *APP* duplications. The genetic tests performed as part of the retrospective study were done in research setting, and were nearly identical to the genetic tests performed as part of the prospective study clinical procedures. There were two exceptions. First, the detection of *APP* duplications was an array-based test in research setting and a Multiplex ligation-dependent probe amplification (MLPA)-based in clinical procedures. Second, we did not evaluate exon-deletions in *PSEN1* in the research setting, while these were evaluated (but not found) in the clinical procedures using MLPA. Complete description of sequencing procedures and MLPA and assessment of *C9ORF72*-repeat length measurement are reported in **Supplementary methods**. Clinical signs sometimes warranted incidental targeted analysis of variants in other genes than those in the panel. These results were extracted from medical records. Clinical laboratory specialist RV and first author SvdL were involved in evaluation of genetic variants for pathogenicity in the research-based retrospective study, while the variants in the prospective study were judged as part of clinical procedures by RV.

### Clinical criteria to determine eligibility for genetic testing

#### Criteria applied (from 2017) until 2021

**Table 1** shows the previous clinical criteria applied to determine eligibility for genetic testing at the Alzheimer Center Amsterdam.^2^

We applied points 1-5 from the criteria to our cohort to identify those patients that would have been eligible for DNA-diagnostics under previous criteria. Note that points 6 and 7, regarding occurrence of disease in family-members, could not be applied because complete family information was not recorded in clinic in 2010-2012. Previously, in our specialized memory clinic only 10-15% of all patients were tested.^2^

#### Design of a novel decision tree and application 2021 onwards

By performing genetic testing, we set out to identify PGV-carriers among all patients who visited the Alzheimer Center Amsterdam between 2010 and July 2012. We used a time window for inclusion to avoid any biases that may be introduced when selecting for age, diagnosis or family history. Based on the number of PGVs identified, we calculated the prevalence of genetic dementia in this retrospective cohort. For each diagnosis group (cognitively normal, mild cognitive impairment [MCI], AD, FTD, etc.), we studied the clinical characteristics of PGV-carriers observed at time of diagnosis. We leveraged these characteristics in the eminence-based design of a decision tree, which reports eligibility for genetic testing and can be more easily implemented in memory clinics than a set of criteria. We aimed to maximize PGV detection by combining the primary diagnosis, age at first presentation and first-degree family history of dementia. Finally, we prospectively applied the decision tree to all patients in clinical care of the Alzheimer Center Amsterdam. We report the number of eligible patients, number of patients tested and PGV detection after one year of clinical implementation.

## Results

In the 2.5-year retrospective study 1022 patients were genetically tested for PGVs. Sex, age at first visit or diagnosis of the 1022 patients who were genetically tested were similar to the 116 patients who were not genetically tested. Of those tested, the average age at first clinical presentation was 62.1 (±8.9SD) years and 413 (40.4%) were female. The tested group had more middle education levels than the not tested group (X^2^ = 1.97, *p* = 0.043, chi-squared test) (**Table 2**).

**Table 2:** Descriptive statistics patients genetically tested vs. those not tested.

| Group | All tested | Not tested | p-value | Statistic |
| --- | --- | --- | --- | --- |
| N | 1022 | 116 |  |  |
| N-females (%) | 413 (40.4%) | 52 (44.8%) | 0.414 | 0.66 |
| Age (years) | 62.1 (8.9) | 62.2 (8.5) | 0.863 | -0.17 (t) |
| Education (low/middle/high) | 4%/58%/38% | 9%/40%/51% | 4.3E-02 | 6.30 |
| *Diagnosis* |  |  |  |  |
| Subjective complaints (%) | 225 (22%) | 18 (15.5%) | 0.134 | 2.25 |
| MCI (%) | 149 (14.6%) | 24 (20.7%) | 0.109 | 2.56 |
| Psychiatry (%) | 104 (10.2%) | 14 (12.1%) | 0.636 | 0.22 |
| AD (%) | 309 (30.2%) | 25 (21.6%) | 0.066 | 3.38 |
| FTD (%) | 51 (5%) | 8 (6.9%) | 0.511 | 0.43 |
| DLB (%) | 35 (3.4%) | 4 (3.4%) | 1.000 | 0 |
| PPA (%) | 3 (0.3%) | 1 (0.9%) | 0.879 | 0.02 |
| Pure vascular dementia (%) | 17 (1.7%) | 5 (4.3%) | 0.108 | 2.58 |
| Other dementias (%) | 30 (2.9%) | 4 (3.4%) | 0.984 | 0 |
| Other neurology (%) | 33 (3.2%) | 2 (1.7%) | 0.545 | 0.36 |
| Unclear diagnosis (%) | 66 (6.5%) | 11 (9.5%) | 0.301 | 1.07 |

Categorical variables were compared using a chi-squared test. Monte Carlo simulation or continuity correction was used if one of the cells in the tables was less than 5. For age an independent t-test was used (t). MCI = mild cognitive impairment, AD = Alzheimer's disease, FTD = frontotemporal dementia, DLB = dementia with lewy-bodies, PPA = primary progressive afasia.

Using the genetic tests, we identified a PGV in 34 out of 1022 (3.3%) patients (**Figure 3A**). The most frequently observed PGV was the hexanucleotide repeat expansion in the *C9ORF72* gene (N = 8, 24% of all PGVs). The second most frequently affected gene was *PSEN1* (NP_000012.1, N=5, p.Ala79Val, 15% of all PGVs), followed by variants in *NOTCH3* (NP_000426.2, N=3, p.Arg1231Cys, p.Arg578Cys, p.Arg640Cys), *SORL1* (NP_003096.1, c.401_402+2delATGT, p.His962Profs*45, p.Pro712Leufs*54), and *TARDBP* (NP_031401.1, p.Ala382Thr[1], p.Asn267Ser[2]) (each gene encompassing, 9% of the total PGVs). We were not aware that patients were related despite patients having the same PGV. One patient had cognitive decline, slowly progressive muscle weakness and spasticity, and a deletion in *SPG4* was identified using MLPA (i.e. outside the dementia gene-panel). One patient was tested for *HTT* repeat expansion because of dementia and subtle chorea: an *HTT*-repeat expansion of 44 repeats was identified (>40 repeats are considered pathogenic). All genes with PGVs are shown in **Table 3**. For privacy reasons, individual patient characteristics and family history of patients with a PGV are not reported.

**Figure 3:**
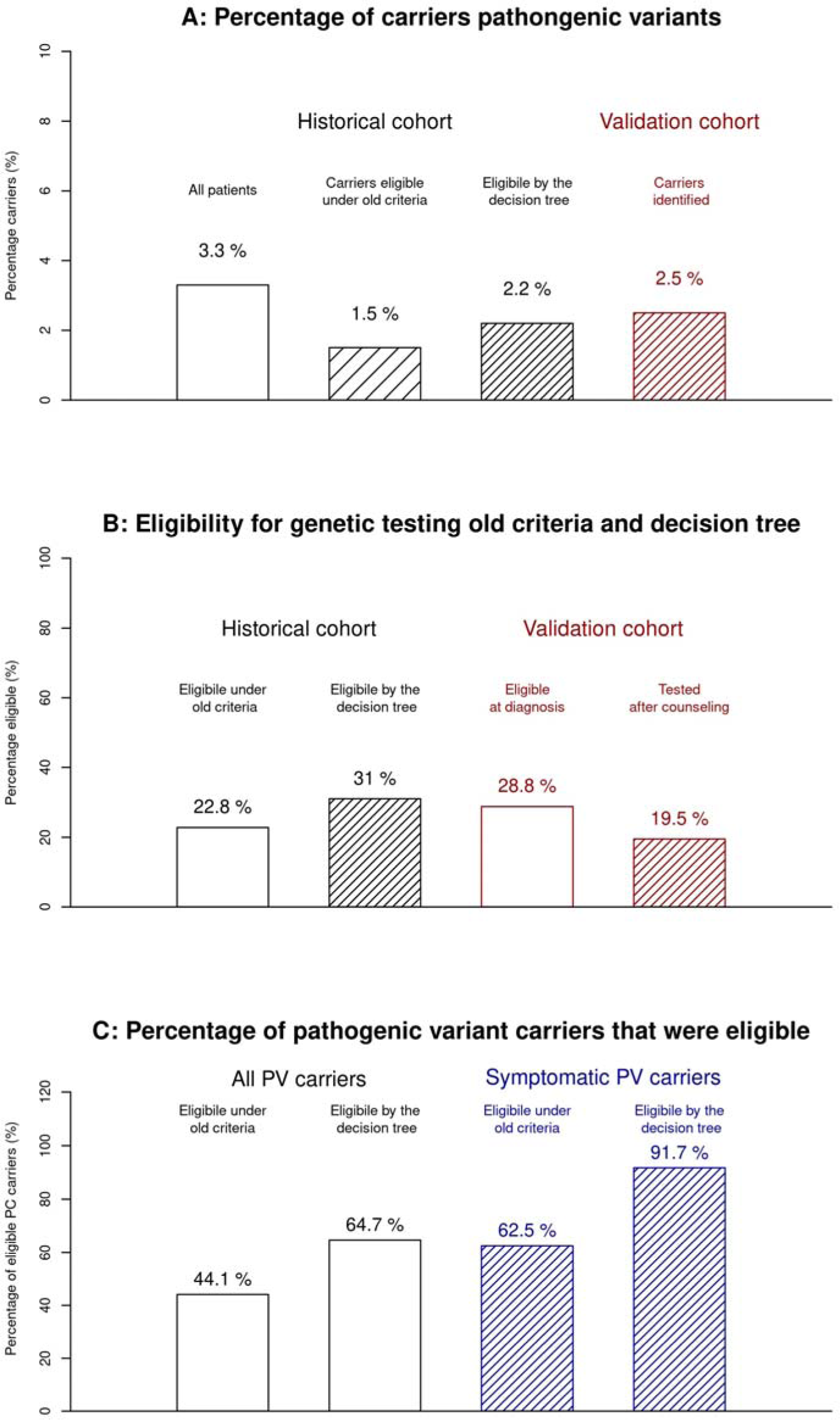
patients are eligible for testing in retrospective and prospective cohort (A), percentage of PGV carriers of all patients in retrospective and prospective cohort (B), number of PGV carriers that were eligible for genetic testing in the retrospective cohort (C).

**Table 3:** genes with PGV in the retrospective cohort

| Gene | N-carriers | Percentage positive | Percentage total |
| --- | --- | --- | --- |
| *C9ORF72* | 8 | 23.5 | 0.8 |
| *PSEN1* | 5 | 14.7 | 0.5 |
| *MAPT* | 3 | 8.8 | 0.3 |
| *NOTCH3* | 3 | 8.8 | 0.3 |
| *SORL1* | 3 | 8.8 | 0.3 |
| *TARDBP* | 3 | 8.8 | 0.3 |
| *CHMP2B* | 2 | 5.9 | 0.2 |
| *CTSF* (homozygote, compound heterozygote) | 2 | 5.9 | 0.2 |
| *APP*-duplication | 1 | 2.9 | 0.1 |
| *GRN* | 1 | 2.9 | 0.1 |
| *Huntington* | 1 | 2.9 | 0.1 |
| *PRNP* | 1 | 2.9 | 0.1 |
| *SGP4* (compound heterozygote) | 1 | 2.9 | 0.1 |
| Negative | 988 | NA | 96.7 |

The demographics of the patients with a PGV were similar to those without a PGV (**Table 4**). The average age at presentation of the 34 patients with a PGV was 58 years (IQR: 52-67), and 62 years (IQR: 56-69) for the 988 patients without a PGV (*p* = 0.056, t-test). For 16 of the 34 patients with a PGV, the age at presentation was <60 (47%), for 13 patients this was between 60 and 70 (38%), and 4 patients were over 70 at presentation (11%).

**Table 4:** Comparing gene carriers to non-carriers

| Group | A PGV present | No PGV | p-value | Statistic |
| --- | --- | --- | --- | --- |
| N | 34 | 988 |  |  |
| N-females (%) | 17 (50%) | 396 (40.1%) | 0.327 | 0.96 |
| Age (years) | 58.9 (9.3) | 62.2 (8.9) | 0.056 | 1.97 (t) |
| Education (low/middle/high) | 0%/59%/41% | 4%/58%/38% | 0.473 | 1.49 |
| *Diagnosis* |  |  |  |  |
| Subjective complaints (%) | 3 (8.8%) | 222 (22.5%) | 0.093 | 2.81 |
| MCI (%) | 4 (11.8%) | 145 (14.7%) | 0.821 | 0.05 |
| Psychiatry (%) | 4 (11.8%) | 100 (10.1%) | 0.982 | 0.00 |
| AD (%) | 8 (23.5%) | 301 (30.5%) | 0.499 | 0.45 |
| FTD (%) | 6 (17.6%) | 45 (4.6%) | 2.3E-03 | 9.28 |
| DLB (%) | 0 (0%) | 35 (3.5%) | 0.524 | 0.40 |
| PPA (%) | 0 (0%) | 3 (0.3%) | 1.000 | 0 |
| Pure vascular dementia (%) | 2 (5.9%) | 15 (1.5%) | 0.203 | 1.62 |
| Other dementias (%) | 1 (2.9%) | 29 (2.9%) | 1.000 | 0 |
| Other neurology (%) | 1 (2.9%) | 32 (3.2%) | 1.000 | 0 |
| Unclear diagnosis (%) | 5 (14.7%) | 61 (6.2%) | 0.102 | 2.67 |

Categorical variables were compared using a chi-squared test. Monte Carlo simulation or continuity correction was used if one of the cells in the tables was less than 5. For age an independent t-test was used (t). MCI = mild cognitive impairment, AD = Alzheimer's disease, FTD = frontotemporal dementia, DLB = dementia with lewy-bodies, PPA = primary progressive afasia.

Of the 34 patients with a PGV, 17 were diagnosed with dementia (50%), subjective memory/cognitive complaints (SMC) (8.8%), mild cognitive impairment (MCI, 11.8%), a primary psychiatric diagnosis (PPD, 11.8%) or an unclear diagnosis (14.7%). When compared by diagnosis, only FTD was overrepresented in PGV-carriers (17.6% vs 4.6%, X^2^=9.2, *p =* 2.3×10^-3^), all other diagnoses were not overrepresented in PGV-carriers. Also, PGV carriers were not more often diagnosed with dementia (50% vs 43%, X^2^ = 0.35, p = 0.55). Surprisingly, only 50% of the PGV-carriers had a reported 1^st^ degree relative with dementia, similar to 49% of the non-carriers (X^2^ = 0, *p* = 1).

### Decision tree for genetic testing in dementia patients (Figure 4)

We constructed a new decision tree to determine eligibility for genetic testing shown in **Figure 4**. First, if a genetic cause is known in the family, direct testing of this PGV is the most logical step (excluding risk factors such as those in the *APOE* gene). If the patient is symptomatic, testing is done by the neurologist as a diagnostic test. If the patient is not symptomatic and interested to know their genetic status, pre-symptomatic counselling by a clinical geneticist is advised **(Figure 4 [1])**.

**Figure 4:**
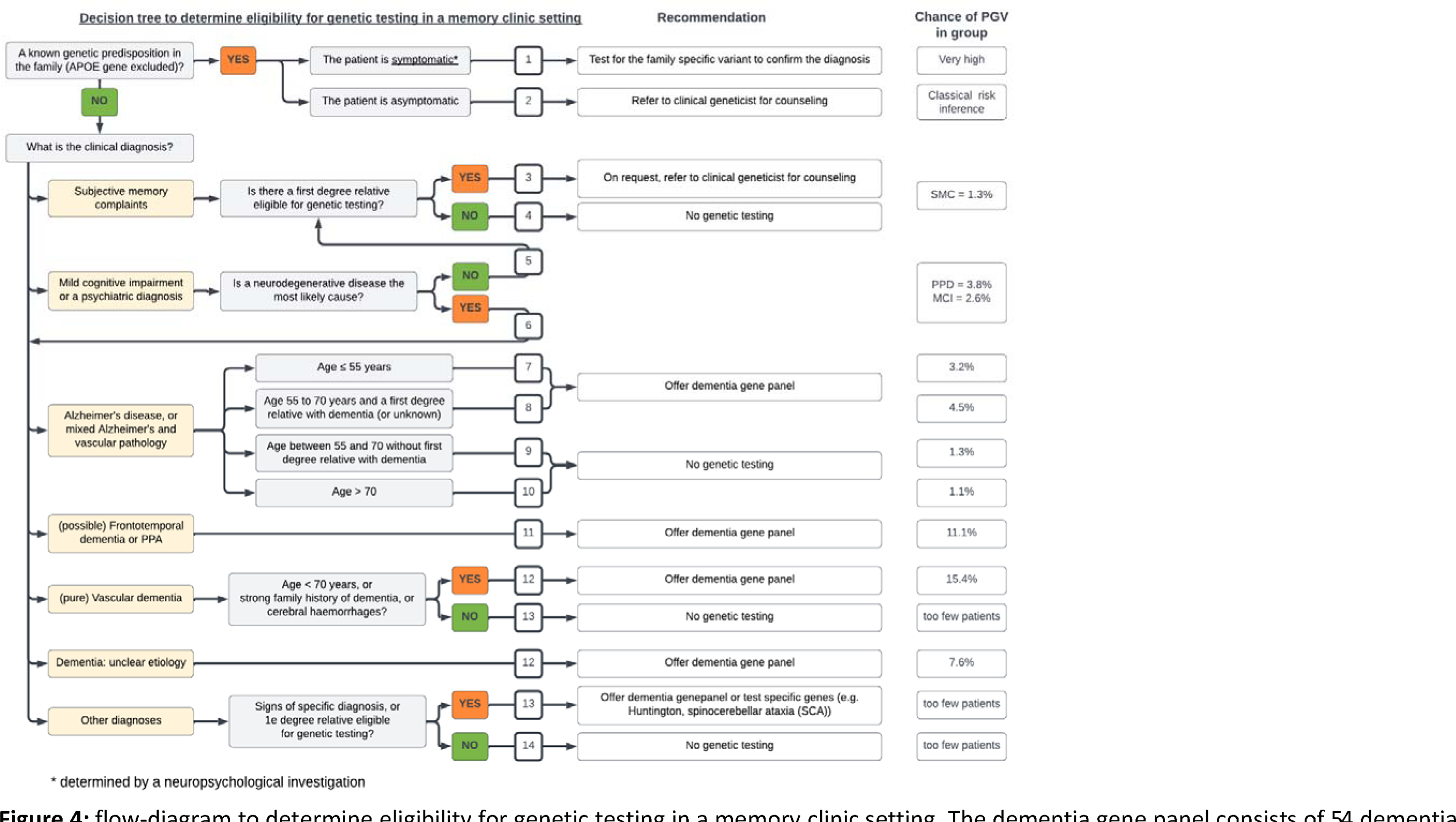
flow-diagram to determine eligibility for genetic testing in a memory clinic setting. The dementia gene panel consists of 54 dementia related genes, the *APP* duplication, and the *C9ORF72* repeat expansion. First degree relatives are: parents, siblings and children. The outcomes of the decision tree are numbered and corresponds to the rows of the table G. The chance to find a pathogenic variant based on the retrospective cohort is shown on the right of the figure.

#### Families with a known PGV

Some patients will have family members were a PGV was previously found. We advise testing of symptomatic patients from such families for the presence of this genetic variant. If a family-member is not (yet) symptomatic and interested in knowing their carrier-ship status, pre-symptomatic counselling is mandatory, and patients should be referred to a clinical geneticist **(Figure 4 [2])**. Subsequently, all remaining steps in the decision tree depend on the clinical diagnosis of the patient.

#### Individuals with subjective memory complaints (SMC)

A diagnosis of SMC is characterized by absence of discernible cognitive decline. We consider all SMC patients as pre-symptomatic and we exercise great care in recommending genetic testing. Still, it is not uncommon for SMC-patients to request genetic testing since SMC can be a very early sign of a neurodegenerative disorder. In patients where a family member meets the criteria for genetic testing, referral to a clinical geneticist for counselling can be considered if patients insist on genetic testing **(Figure 4 [3])**. In our cohort, 3 out of 225 (1.3%) SMC patients were PGV carriers. One patient was identified to carried the pathogenic repeat expansion in C9ORF72 and was diagnosed with behavioural variant of FTD 10 years after initial presentation. The two patients were discharged and no further information was available.

#### Individuals with primary psychiatric diagnosis (PPD) and minor cognitive impairment (MCI)

A diagnosis of PPD and MCI can be the first measurable effect of a neurodegenerative disease, but MCI may also occur due to other non-neurodegenerative causes such as a burn-out or sleep disturbances. Therefore, the decision tree includes an intermediate step for patients with MCI or PPD. For these patients, the clinician has to determine whether the underlying cause of MCI or PPD is most likely a neurodegenerative disease. Examples when genetic testing could be considered are: a very strong family history, MCI and a positive test for AD-biomarkers in CSF (or PET-scan), or PPD with signs of neurodegeneration on brain MRI. In the retrospective study, 4 of the 104 PPD patients carried a PGV (3.8%). In the follow-up of these PGV-carriers one patient was identified as PGV carrier in routine clinical care. We found a MAPT PGV in a patient with a depression. In the follow-up of this patient a family member developed FTD, was genetically tested and carried the same PGV in *MAPT* that was also identified in our patient. Our patient eventually underwent pre-symptomatic counselling and the *MAPT* variant was disclosed. The patient is still under yearly evaluations and we have not observed progression.

Among the patients with MCI (N = 149), we identified 4 patients with a PGV (2.6%). In two patients, we identified a PGV in the *CHMP2B* gene. In one patient, we observed slow progressive decline over 8 years and development of muscle weakness. The second MCI patient carried a PGV in *NOTCH3*. This patient was diagnosed with MCI and, at first follow up, the patient progressed to vascular dementia followed by a fast clinical decline and death. The last MCI patient, had positive amyloid biomarkers in cerebrospinal fluid and carried a PGV in *PSEN1*. Taken together we recommend to exercise care in recommending genetic testing in patients with MCI and psychiatric **(Figure 4 [5]).** Only if a neurodegenerative disease is considered as the plausible explanation for their complaints, patients with MCI and psychiatric disease can be considered for genetic testing and the decision can be based on the most likely underlying neurodegenerative disease **(Figure 4 [6])**.

#### Alzheimer’s Disease (AD)

We recommend genetic testing in all patients with very early onset AD. In the 31 patients with a very young age at presentation (≤55years). We identified a duplication of the *APP* gene in one patient with very early onset AD (3.2%) **(Figure 4 [7])**. We included family history (or unknown family history) into the decision tree for the AD patients aged between 55 to 70 years, as those with a positive family history or unknown family history were more likely to be a PGV carrier: in 5 of the 110 AD patients (4.5%) with a 1^st^ degree relative with dementia we observed a PGV **(Figure 4 [8])**. In contrast, we observed a PGV in only 1 out of 77 AD patients (1.1%) without a first degree relative **(Figure 4 [9])**. In patients aged over 70 years we do not recommend genetic testing **(Figure 4 [10]).** In the 91 patients with first presentation at age >70, we observed a PGV in *NOTCH3* in an a patient with AD (p.Arg1231Cys) (1.1%). In the differential diagnosis of this patient, vascular dementia was considered, it is unclear what the primary pathology is in this patient.

#### Frontotemporal dementia (FTD) or PPA

Clinical guidelines recommend genetic testing for patients with FTD and PPA. We therefore included this recommendation in diagnostic decision tree. Indeed, among the 54 patients diagnosed with FTD, we identified 6 PGV carriers (11.1%). **(Figure 4 [11])**.

#### Vascular dementia

In other memory clinics with mainly older patients, vascular dementia is mostly due to vascular risk factors such as hypertension and stroke. We therefore put a separate step in the decision tree, in which we advise that only patients with vascular dementia with an age at presentation <70 years and/or a 1^st^ degree relative with dementia be eligible for genetic testing **(Figure 4[12 & 13])**. In our data, 13 patients fulfilled these criteria and were diagnosed with ‘vascular dementia, of which 2 carried a PGV (15.4%) in and a *C9ORF72* repeat expansion.

#### Uncertain dementia types

We recommend testing in patients with uncertain type of dementia. Of the 66 patients for whom the subtype of dementia remained uncertain, 5 patients (7.6%) were affected by PGVs in respectively the *HTT, C9ORF72, GRN, CTSF, TARDBP* genes **(Figure 4 [14])**.

#### Other dementias

This group includes, for example, dementia with *Lewy Bodies (DLB).* Among the 35 patients diagnosed with DLB we identified no PGV carriers. Therefore, we exercise caution in recommending genetic testing for DLB cases. However, when there is a strong family history, or when neurological symptoms indicative of rare genetic neurological disorders manifest (such as chorea in Huntington’s disease or ataxia in spinocerebellar ataxia), a gene panel and targeted test for the specific PGVs or a gene associated with these rare neurological disorders is advised **(Figure 4 [15 & 16])**.

### Retrospective application of old and revised criteria

The retrospective study was conducted on 1,022 patients that entered the ADC between 2010 to 2012. The research database of the ADC contained the information to apply the old clinical criteria to determine if patients were eligible for genetic testing (**Table 1**). This allowed us to compare the identification of PGV carriers between the old and the new decision tree (**Table 1**). Based on the old clinical criteria, we estimate that 261 of 1,022 patients would have been eligible for genetic testing (22.8%) (**Figure 3B**), allowing the identification of only 15 PGV-carriers (15/34 = 44.1%) while 19 PGV-carriers would have been missed (19/34 = 56%) (**Figure 3C**). Based on the newly formulated eligibility criteria, we estimate that 317 (31.0%) of the 1,022 patients present with characteristics that would have rendered them eligible for testing (**Figure 3B**) which would have led to the identification of 22 out of 34 PGV-carriers, an identification rate of 65%. This is 2.2% of the retrospective cohort (**Figure 3 C**). When we recalculated the eligibility for symptomatic patients under the old and new criteria (excluding patients with subjective memory complains, psychiatry or MCI with amyloid biomarkers), a total sample of 606 (59%) remain. In this sample, 24 individuals have a PGV (24/606 = 3.9%), from which under the old criteria 15 (15/24 = 62.5%) would have been eligible and with the decision tree 22 patients with a PGV would be eligible (22/24 = 91.7%).

### Prospective Application of the revised criteria in a memory clinic (validation)

We prospectively applied the decision tree in all patients entering the Alzheimer Center Amsterdam for one year. The clinical work-flow was adapted as shown in **Figure 2**. We implemented four main adaptations to the previous workflow to accommodate this. In practice, the medical record, MRI and family history of a patient were discussed prior to the multidisciplinary meeting by the staff of the clinical genetics department. In our prospective application this was done by a research physician (SvdL) and clinical geneticist (CG, MWE). In this pre-discussion, it was decided if there was an indication for genetic testing, and, if so, whether there was a high suspicion of monogenic dementia based on family history. At the multidisciplinary meeting of the memory clinic this outcome would be shared, and if there was high suspicion, a referral for counselling, this would be recommended to the treating neurologists. In some instances, the advice was modified based on new insights revealed at the meeting. For the neurologist, the most practical moment to offer genetic testing or counselling was at the patient-visit at which the diagnosis was communicated to the patient. The neurologist informed whether the patient was eligible for genetic testing, or if referral was indicated. In this consultation with the neurologist, patients decided either; a) not to opt for genetic testing, b) opt for genetic testing, or c) to be referred to a clinical geneticist for counselling in decision-making. In some instances, the neurologist postponed the decision to a follow-up visit.

In total, 97% (517/533) of patients who visited the clinic between September 2021 and September 2022 were entered in the prospective cohort. The average age at presentation was 64.1 years (SD 8.5) and 41.6% of patients was female. The results of the validation cohort are shown in **Figure 5** and **Table 5**. Two patients had already undergone genetic testing prior to their visit (2/517= 0.4%). Of the remaining 515 patients, a total of 148 patients (28.6%) were eligible for genetic testing at time of diagnosis. This was comparable to the estimation based on the retrospective cohort in this clinic (31%). Four months after the last patient was included in the prospective cohort, 103 patients had consented to genetic testing (103/517=20%) of whom, at the multidisciplinary meeting, 90 were considered eligible according to the decision tree (90/103=88%). In 13 patients who were not deemed eligible for genetic testing by the criteria, the neurologist decided to go ahead with genetic testing regardless. In 13 out of the 103 patients that were tested a PGV was found. This corresponds to a diagnostic yield of 12.6% (13/103) and 2.5% a minimal prevalence of genetic dementia in the total cohort (13/515, **Figure 3A**). Based on the prevalence of monogenic dementia in the retrospective cohort (3.3%), we expected ∼17 PGV in the prospective cohort (3.3% of 515 patients). With the old criteria 44% of 17 PGVs would have been expected to be identified (7.48 patients). We therefore estimate that we identified 73% more PGV than expected (13/7.48) and 76% (13/17) of the anticipated PGVs.

**Figure 5:**
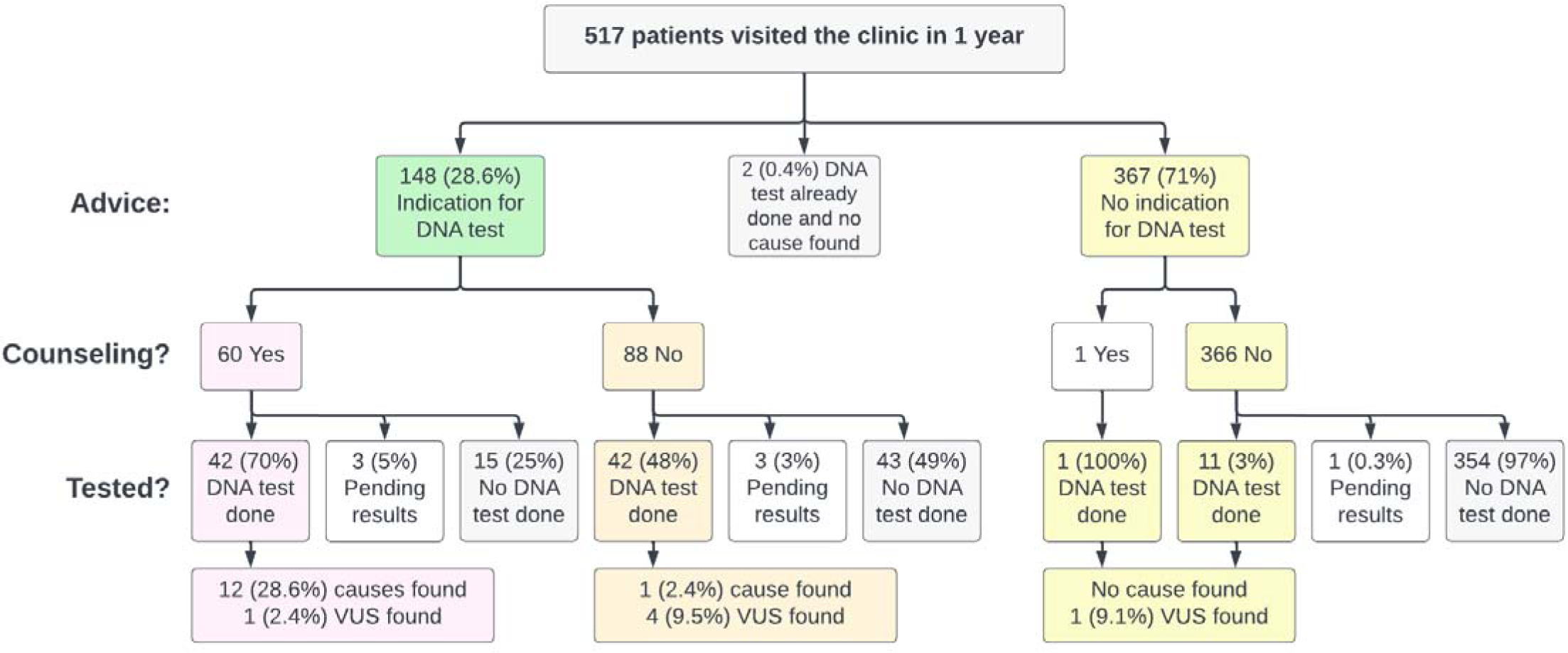
results of 1 year prospective implementation in clinical care.

**Table 5:** Results of the validation cohort

| Flow-chart outcome |  | Advice from decision tree | N-patients (% of total) | N-eligible (% of group) | N-tested (% of eligible) | N-carrier (% of tested) |
| --- | --- | --- | --- | --- | --- | --- |
| 1 | PGV known: symptomatic | Test for PGV | 1 (0.2) | 1 (100) | 1 (100) | 1 (100) |
| 2 | PGV known: a-symptomatic | Refer | 2 (0.4) | 2 (100) | 1 (50) | 1 (100) |
| 3 | SCD/psychiatry/MCI: test indicated | Refer | 5 (1) | 5 (100) | 3 (60) | 2 (66.7) |
| 4 | SCD/psychiatry/MCI: test not indicated | No test | 169 (32.7) | 0 | 0 | 0 |
| 5 | AD: <55 years of age | Offer test | 16 (3.1) | 16 (100) | 5 (31.2) | 0 |
| 6 | AD: 55-70 years of age with 1e degree relative with dementia | Offer test | 52 (10.1) | 52 (100) | 23 (44.2) | 0 |
| 7 | AD: 55-70 years of age test no 1e degree relative with dementia | No test | 59 (11.4) | 3 (5.1) | 6 (200)* | 0 |
| 8 | AD: >70: test not indicated | No test | 61 (11.8) | 0 | 2 | 0 |
| 9 | FTD/PPA: test indicated | Offer test | 21 (4.1) | 20 (95.2) | 16 (80) | 1 (6.2) |
| 10 | Vascular dementia: age<70 or family history or cerebral haemorrhage | Offer test | 7 (1.4) | 7 (100) | 4 (57.1) | 3 (75) |
| 11 | Vascular dementia other | No test | 2 (0.4) | 0 (0) | 0 | 0 |
| 12 | Unclear aetiology | Offer test | 35 (6.8) | 34 (97.1) | 27 (79.4) | 3 (11.1) |
| 13 | Other diagnoses: signs of specific cause or 1e degree relative fulfilling eligibility criteria | Offer test or test specific genes | 7 (1.4) | 7 (100) | 6 (85.7) | 2 (33.3) |
| 14 | Other diagnoses: test not indicated | No test | 49 (9.5) | 1 (2) | 3 (300)* | 0 (0) |

The outcomes of the decision tree in figure 3 are numbered and corresponds to the rows of the table. N- eligible is the number of patients determined eligible for genetic testing. \*The percentage of patients tested is higher than the number of patients eligible as neurologists performed DNA-tests in patients not eligible. Possibly due to changes in clinical symptoms, family history, diagnosis or on patients' request.

**Table 6:** Genes and mutations in the validation cohort.

| Gene | Mutation(s) | N-carriers | Percentage of PGVs | Percentage total |
| --- | --- | --- | --- | --- |
| *NOTCH3* | NM_000435.2(NOTCH3):c.1732C>T p.(Arg578Cys) heterozygote | 3 | 23.1 | 0.6 |
|  | NM_000435.2(NOTCH3):c.2182C>T p.(Arg728Cys) heterozygote |  |  |  |
|  | NM_000435.2 (NOTCH3):c.3910T>C p.(Cys1304Arg) heterozygote |  |  |  |
| *PSEN1* | PSEN1(NM_000021.3):c.1254G>C p.(Leu418Phe) heterozygote | 1 | 7.7 | 0.2 |
| *PSEN2* | NM_000447.3(PSEN2):c.715A>G p.(Met239Val) heterozygote | 2 | 15.4 | 0.4 |
|  | NM_000447.2(PSEN2):c.766del p.(Leu256Trpfs*19) heterozygoot |  |  |  |
| *MAPT* | NM_005910.5 (MAPT):c.1216C>T p.(Arg406Trp) heterozygote | 1 | 7.7 | 0.2 |
| *HTRA1* | NM_002775.4(HTRA1):c.961G>C p.(Ala321Pro) heterozygote | 1 | 7.7 | 0.2 |
| *CTSF* | NM_003793.3(CTSF):c.965-1G>A p.? homozygote | 2 (siblings) | 15.4 | 0.2 |
| *AMACR* | NM_014324.6(AMACR):c.155C>A (p.Ser52*), c.1040dup (p.Glu348Argfs*4) Compound heterozygosity | 1 | 7.7 | 0.2 |
| *AARS2* | NM_020745.3(AARS2):c.2255+1G>A p.?, c.372C>A p.(Asp124Glu) Compound heterozygosity | 1 | 7.7 | 0.4 |
| *duplication of 16q24.2q24.3* | 16q24.2q24.3(88630655_89772750)x3 | 1 | 7.7 | 0.2 |
| Negative |  | 506 |  | 97.9 |

PGVs were observed in *PSEN1* (1x), *PSEN2* (2x), *NOTCH3* (3x), *MAPT* (1x), *HTRA1*(1x), *CTSF* (2x compound heterozygote), *AMACR* (1x compound heterozygote, specific test), *AARS2* (2x compound heterozygote), and a duplication in the region 16q24.2q24.3 (specific test performed because of concomitant intellectual disability) (**Table 5**). It is noteworthy that we did not identify any carriers of the FTD/ALS associated *C9ORF72* repeat expansion.

#### Variants of unknown significance (VUS)

We observed a VUS in 6 out of 103 patients (5.8%). In the families of carriers, no additional segregation analysis or functional analysis could be done in family members of the patients with a VUS, and follow-up by clinical geneticists did not alter the classification of the VUS. We recommend revision of these genetic variants after 5 to 10 years.

#### Implementation of counselling

In our patient cohort, 60 out of 148 patients who were eligible for genetic testing were counselled by a clinical geneticist (40.5%), and 88 were counselled by the neurologist (59.5%). After counselling by clinical geneticist 45 (75%) agreed to proceed with testing, while 15 patients decided against testing (25%). After counselling by clinical neurologist 45 (51%) proceeded with testing, while 43 patients decided against testing (49%). Of the 42 patients that had completed a genetic test after counselling by the clinical geneticist 26.6% (12/42) carried a PGV. In contrast, of the 42 patients that completed a genetic test after counselling by the neurologist 2.4% (1/42) carried a PGV. This is in line with referral of high suspicion patients for counselling by a clinical geneticist prior to testing.

## Discussion

Offering genetic tests to the right patients visiting specialized memory clinics can lead to identification of PGVs, which are crucial for explaining disease causation, facilitating clinical trial participation, enabling pre-symptomatic testing in healthy relatives, and informing preventive measures in family planning. We found that based on previous criteria only 65% of all symptomatic patients with a PGV were not eligible for genetic testing and developed a decision tree that increased this to 91% of symptomatic patients with a PGV. We prospectively implemented the decision tree, which resulted in 73% more than expected identified PGVs. Our novel decision tree can aid clinicians in memory clinics to select patients eligible for genetic testing.

In literature there is one study, that also searched for monogenic causes of dementia in a large number of samples. This study by Koriath *et al*. investigated a substantial referral-based dementia cohort encompassing AD, FTD, and prion diseases, and found deleterious variants in 12.6% of all patients using a targeted dementia gene panel consisting of 17 genes (along with the *C9orf72* expansion and *PRNP* octapeptide repeat alteration)^20^. In our single-site clinical cohort within a specialized memory clinic, the prevalence of PGVs was much lower (3.3%). This is a seemingly large difference, but the patient population of our memory clinic was very different from the referral-based cohort. For example, Koriath *et al*. itincluded 24.4% FTD patients (compared to 5% in our study) and 9.2% had a prion disease (compared to 0.2% in our study grouped under other diagnoses).^14^ The number of PGV carriers in these groups were very high >20%, increasing the total percentage of patients with a PGV. Also, the number of patients without cognitive complaints (controls and PPD) was higher in our study (32%) compared to Koriath *et al*. (14%).^14^ In both studies the percentage of controls with a PGV was low, therefore decreasing our percentage of carriers. If we match the percentage of disease prevalences of Koriath *et al.* to our population, the percentage of PGV carriers in Koriath *et al.* would be between 4 and 5%, which is only slightly higher than our estimate.

The clinical criteria for genetic testing in dementia patients worldwide are variable. Typically, these criteria involve a combination of a clear clinical phenotype, a (very) early-onset of complaints, or a strong family history of dementia. In previous studies this resulted in diagrams of varying complexity,^2,6,14,20,22,32–34^ often for separate diseases, such as AD^33^, MND-FTD, HTT and prion diseases,^14^ or criteria based on well-defined patient cohorts,^14^ and in cases of unclear etiology.^35^ Our decision tree resembles most the procedure for eligibility presented by Koriath *et al.*^20^, but we put less emphasis on family history and used age at presentation. To increase acceptance and practical use of the decision tree, we used the clinical diagnosis at presentation, and only require family history of dementia of 1^st^ degree relatives. We used the age at presentation instead of age at onset as it is easier for the clinician and is not influenced by recall bias. The age cut-offs implemented are also much less strict than, for example, the NHS criteria where an age at onset of less than 55 years is advised.^23^ Less emphasis on a very early onset is necessary as we found that 53% of patients with a PGV was over 60 at time of presentation, replicating similar previous findings.^14,36^ For family history the most used criteria are the Goldman criteria for genetic testing for patients with FTD^34^ and AD^21^. In daily clinical practice the diagnosis may not be clear, and an in-depth family history may not be present, or even absent.^20^ Still, if detailed family information is present clinicians should consider the wider family history.^6,14,34^ In our validation cohort, we used family history to determine if counselling should be done by a clinical geneticist or the neurologist. Based on our observations, we suggest to make referral to a geneticist a perquisite before initiating a genetic testing when there is strong family history of dementia, unexplained psychiatry.

Patient and family perspective is the most important consideration when genetic testing is offered. The general interest in genetics is rising because of an increasing tendency to diagnose patients with dementia earlier in their disease course^37^, options to participation in clinical trials^10,11^, the prospect of disease modifying therapies, and to arrange life decisions for patients with young onset dementia. Not finding a PGV could bring relief, and finding a PGV can be of high impact. The consequences of finding more PGV carriers on patients and family members will have to be evaluated in clinical care.

This study has several strengths that enhance its reliability and validity. First, embedding in the Alzheimer Center Amsterdam provided a significant advantage, as it ensured a uniform and streamlined workflow for all patients.^30^ This consistency and accuracy in the clinical assessments of patients improved the reliability of reported numbers of expected patients with PGV by diagnosis. Second, the study utilized a large clinical sample, reflecting a real-world clinical care setting. The prospective implementation of the decision tree further strengthens the validity of the findings. Third, approximately 95% of all patients entering the clinic participate in scientific research, minimizing bias related to age, diagnosis, and clinical center. However, it is essential to note that the Alzheimer Center Amsterdam specializes in young onset dementia with many patients asking for a second opinion. This likely led to a higher percentage of eligibility and a larger number of carriers of PGV compared to an average memory clinic. While this was crucial for constructing and validating the decision tree, it also limited the number of patients over the age of 75 years. External studies have shown lower numbers of patients with deleterious variants in this age group.^14^ Further validation in less specialized centers with older patients and fewer additional diagnostic tests is warranted. An additional challenge in genetic testing is the identification of gene variants of unknown significance. It is a limitation that we did not systematically assess the number of uncertain significance (VUS, class 3 variants) in our retrospective cohort. Still, in the prospective cohort the number of observed variants of VUS was low (5.8%) and had no clinical consequences. Last, despite the study’s use of a panel of 54 genes, we acknowledge the ever-expanding list of genes associated with dementia since the study’s initiation in 2017. Our targeted gene panel was updated at the end of the study, and we advise using this updated panel for genetic testing (**Supplementary table 2**).

For policy makers and care organisation it is essential to have an estimated number of patients eligible for genetic testing. Based on our results, we estimate that approximately one in five patients below the age of 70 years, regardless of their specific diagnosis, would be eligible for testing and would want to be tested. We estimate, based on our prospective study, that 38% of the patients tested will need consultations by a clinical geneticist or trained physician. Between the ages of 70 and 75, approximately one in twenty patients would be tested and (nearly) no patients over the age of 75. It is important to consider that these numbers will be lower in less specialized memory clinics and different healthcare systems. Restricted financial options in different health care systems, and varying legal protections in different countries, may also influence patients’ decisions regarding genetic testing.

In conclusion, routine assessment of eligibility for genetic testing in all patients with dementia will improve the identification of patients with monogenic dementia. Implementation requires minor change from ‘standard’ clinical practice and with the decision tree clinicians can offer genetic testing to the right patients, leading to more personalized management options of patients with dementia.

## Data Availability

All data produced in the present study are available upon reasonable request to the authors

## Acknowledgements

We thank all study participants and all personnel involved in data collection for the contributing studies. SvdL was funded for this study by NWO (#733050512, PROMO-GENODE: a PROspective study of MOnoGEnic causes Of Dementia) a substantial donation by Edwin Bouw Fonds and Dioraphte. SvdL further received funding for the GeneMINDS consortium, which is powered by Health∼Holland, Top Sector Life Sciences & Health. Research of Alzheimer Center Amsterdam is part of the neurodegeneration research program of Amsterdam Neuroscience. Alzheimer Center Amsterdam is supported by Stichting Alzheimer Nederland and Stichting Steun Alzheimercentrum Amsterdam. The chair of Wiesje van der Flier is supported by the Pasman stichting. The clinical database structure was developed with funding from Stichting Dioraphte. SvdL, HH, MR, WF are recipients of ABOARD, which is a public-private partnership receiving funding from ZonMW (#73305095007) and Health∼Holland, Topsector Life Sciences & Health (PPP-allowance; #LSHM20106). More than 30 partners participate in ABOARD. ABOARD also receives funding from Edwin Bouw Fonds and Gieskes-Strijbisfonds. ABOARD also receives funding from de Hersenstichting, Edwin Bouw Fonds and Gieskes-Strijbisfonds. Array genotyping was performed in the context of EADB (European Alzheimer DNA biobank) funded by the JPco-fuND FP-829-029 (ZonMW projectnumber 733051061). IdR is supported by the ISCIII under the grant FI20/00215.

The work in this manuscript was carried out on the Cartesius supercomputer, which is embedded in the Dutch national e-infrastructure with the support of SURF Cooperative. Computing hours were granted in 2016, 2017, 2018 and 2019 to H. Holstege by the Dutch Research Council (project name: ‘100plus’; project numbers 15318 and 17232).

