## Supplementary Methods for "Genetic Testing in patients with Dementia: A Data-Driven Clinical Decision Tree for Memory Clinics"

### Supplementary methods and figures:

### Content of genetic test

#### 1. Dementia gene-panel.

We generated whole exome sequencing data using the Agilent v4 or v6 kits (~58Mb target region). Sequencing was done on Illumina NovaSeq 6000 (2x150basepair reads) and samples had at least 8Gb raw data per sample. A uniform pipeline was used to process all samples as previously described.[31] Raw sequencing data from all studies were processed relative to the GRCh37 reference genome, the read alignments of possible chimeric origin were filtered and a GATK-based pipeline was used to call variants, while correcting for estimated sample contamination percentages. Samples were included in the datasets after they passed a stringent quality control (QC) pipeline: samples were removed when they had high missingness, high contamination, a discordant genetic sex annotation, high numbers of new variants (with reference to dbSNP v.150), deviating heterozygous/homozygous or transition/transversion ratios. We did not exclude patients with non-european ancestry.[31] In total 11 patients could not be tested due to sequencing data failing these QC steps. We extracted genetic variants in the exons and flanking regions (+/- 6 basepairs) from 54 genes associated with monogenic dementia (Supplementary table 1) for each patient. All variants were annotated for pathogenicity in the Alissa Interpret® software (Agilent Technologies - v5.4.0). Variant classification followed the guidelines published by the American College of Medical Genetics and Genomics and the Association for Molecular Pathology.[20] The classification is based on the level of evidence available for each variant. Intronic variants were assessed using webtool SpliceAI (https://spliceailookup.broadinstitute.org/). Only variants with population frequencies <1% were considered. In the retrospective study, variants that were unclassified after initial filtering steps based on frequency, were classified independently by a trained researcher (SvdL) and a clinical molecular geneticist (RV). Variants that were classified by one or both as class III/IV/V were discussed, a consensus was reached and only class IV/V were considered pathogenic variants . In the prospective study only a clinical molecular geneticist (RV) interpreted the variants for pathogenicity.

#### 2. *Assessing C9orf72* hexanucleotide repeat expansions

We used the standard methods described in Renton et al.[25] with minor modifications[32] to measure the *C9orf72* repeat lengths. All expansions, and if only 1 allele was present, were followed-up by either repeat-primed PCR or a commercial kit (AmplideX PCR/CE C9orf72 Kit, Asuragen). (Reus et al., 2021). Hexanucleotide repeat lengths over 30 were considered pathogenic. In the implementation cohort *C9orf72* repeat lengths were determined with a commercial kit (AmplideX PCR/CE C9orf72 Kit, Asuragen).

#### 3. Assessing APP duplications

In the retrospective cohort patients were genotyped on the Illumina® genome screening array (GSA) a high density single nucleotide polymorphism (SNP)[33], Illumina® BeadStudio was used to extract the the log R ratio (LRR) and B allele frequency (BAF) per individuals for all genotyped SNPs on chromosome 21. To determine the presence of CNVs, we used the PennCNV software[34]. PennCNV uses a Hidden Markov Model (HMM), in which it incorporates the LRR and BAF[35]. The combination of these two was used to determine the CNVs. We followed the user guidelines of PennCNV [34] and extracted all duplications covering the full *APP* gene (chr 21:27252861-27543446, HG build 37). In the prospective cohort *APP* duplications and *PSEN1* deletions were measured with commercial MLPA- Multiplex Ligation-Dependent Probe Amplification (MLPA) (P170-C3 and P254-B2, MRC Holland).

**Supplementary table 1:** genes in dementia gene panel used in current work

Hexanucleotide C9ORF72 repeat, ALS2 (NM_020919.3), ANG (NM_001145.4), APOE (NM_001302688.1), APP (NM_000484.3), duplications of APP(NM_000484.3), ATP7B (NM_000053.3), C19ORF12 (NM_001031726.3), C9ORF72 (NM_001256054.2), CHCHD10 (NM_001301339.1), CHMP2B (NM_014043.3), CLN3 (NM_001042432.1), CLN5 (NM_006493.2), CP (NM_000096.3), CSF1R (NM_005211.3), CTSD (NM_001909.4), CTSF (NM_003793.3), EIF4G1 (NM_182917.4), ERBB4 (NM_005235.2) FUS (NM_004960.3), GRN (NM_002087.3), HNRNPA1 (NM_031157.3), HNRNPA2B1 (NM_031243.2), HTRA1 (NM_002775.4), ITM2B (NM_021999.4), MAPT (NM_005910.5), NOTCH3 (NM_000435.2), NPC1 (NM_000271.4), NPC2 (NM_006432.3), OPTN (NM_001008211.1), PDGFB (NM_002608.3), PDGFRB (NM_002609.3), PPT1 (NM_000310.3), PRKAR1B (NM_001164761.1), PRNP (NM_000311.3), PSEN1 (NM_000021.3), PSEN2 (NM_000447.2), PSENEN (NM_172341.3), SERPINI1 (NM_005025.4), SETX (NM_015046.5), SIGMAR1 (NM_005866.3), SLC20A2 (NM_001257180.1), SNCA (NM_000345.3), SNCB (NM_001001502.2), SOD1 (NM_000454.4), SORL1 (NM_003105.5), SPG11 (NM_025137.3), SQSTM1 (NM_003900.4), TARDBP (NM_007375.3), TBK1 (NM_013254.3), TREM2 (NM_018965.3), TYROBP (NM_003332.3), UBQLN2 (NM_013444.3), VCP (NM_007126.3), VPS13A (NM_033305.2), XPR1 (NM_004736.3).

**Supplementary table 2:** Genes in updated panel (2022)

duplications of APP(NM_000484.3), hexanucleotide C9ORF72 repeat. AARS2, ABCD1, ADAR, APP, ARSA, ASPA, C19ORF12, C9ORF72, CCNF, CHCHD10, CHMP2B, CLCN2, CLN3, CLN5, CLN6, COL4A1, COL4A2, CP, CSF1R, CTSA, CTSD, CTSF, CYLD, DARS2, DCTN1, DNAJC5, DNAJC6, DPP6, EIF2B1, EIF2B2, EIF2B3, EIF2B4, EIF2B5, EPM2A, ERBB4, FA2H, FUS, GALC, GBE1, GNS, GRN, HEXA, HGSNAT, HMBS, HTRA1, IDS, IDUA, IFIH1, ITM2B, JAM2, LMNB1, MAPT, MATR3, MCOLN1, MYORG, NAGLU, NHLRC1, NOTCH3, NPC1, NPC2, OPTN, PANK2, PDGFB, PDGFRB, PINK1, PLA2G6, PPT1, PRKAR1B, PRNP, PSAP, PSEN1, PSEN2, RNASEH2A, RNASEH2B, RNASEH2C, RNASET2, SAMD9, SAMD9L, SAMHD1, SERPINI1, SGSH, SLC20A2, SMPD1, SNCA, SNCB, SPG11, SQSTM1, STUB1, TARDBP, TBK1, TMEM106B, TREX1, TUBA4A, TYROBP, UBQLN2, VCP, VPS13A, XPR1.

### Supplementary figure without the numbers

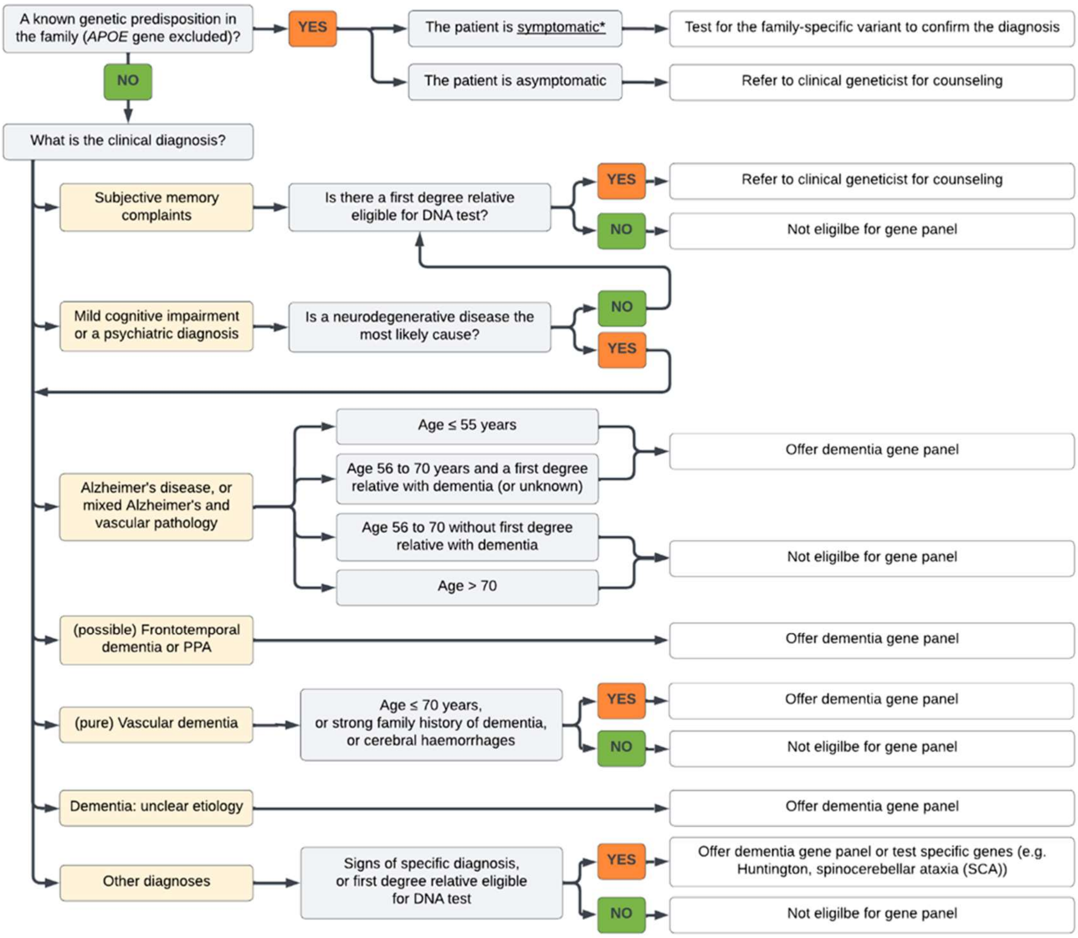

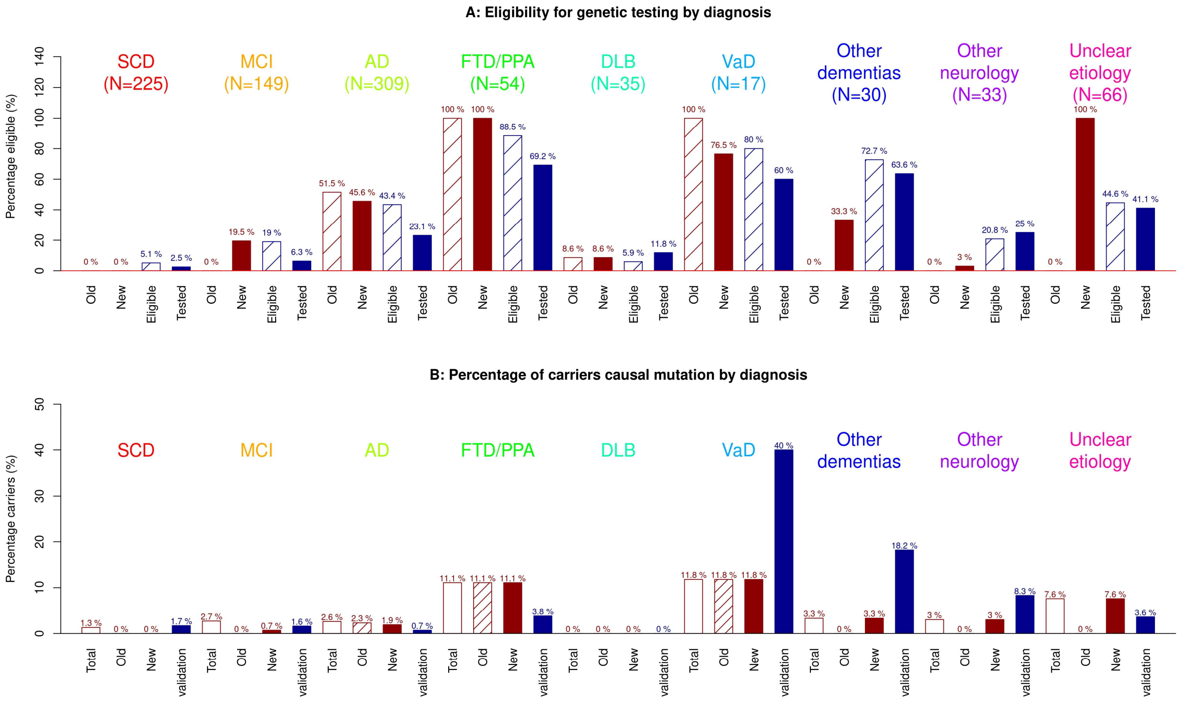


**Supplementary Fig. 2A:** Distribution eligibility by diagnosis in the historical data using the old criteria (red dashed), historical data using the new criteria (red filled), the eligibility prospective cohort (blue dashed) and percentage of eventually tested patients in the prospective cohort (blue filled). Note that the percentage of 'eligible' may be lower than expected in the validation cohort as clinincians may have decided to deviate from the decision tree. **2B**: percentage of carriers of a pathogenic genetic variant (PGV) by diagnosis. Total (red, empty) = percentage of carriers in all patients of the historical cohort, old (red, dashed) = percentage of carriers in those eligible according to old criteria in the historical cohort, new (red, filled) = percentage of carriers eligible according to the new criteria in the historical cohort, validation (blue filled) = percentage of carriers with a PGV in the prospective cohort.

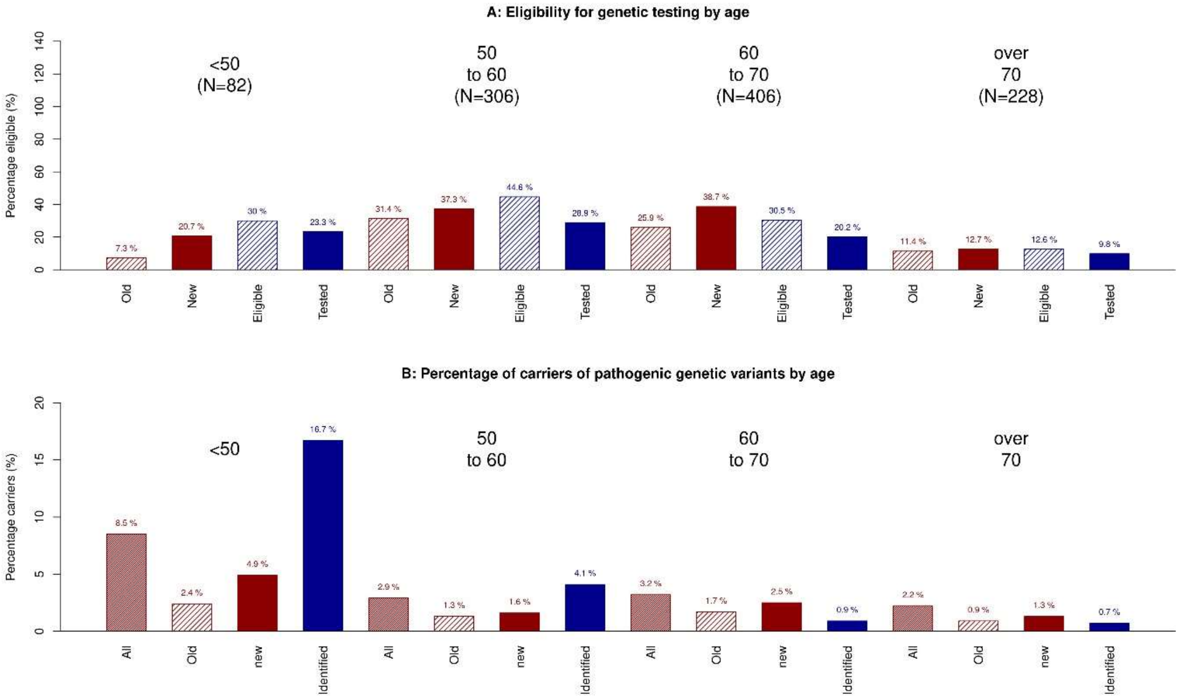


**Supplementary Fig.3:** Eligibility and percentage of carriers pathogenic genetic variants by Age in the retrospective cohort and prospective implementation cohort. Total (red, empty) = percentage of carriers in all patients of the historical cohort, old (red, dashed) = percentage of carriers in those eligible according to old criteria in the historical cohort, new (red, filled) = percentage of carriers eligible according to the new criteria in the historical cohort, validation (blue filled) = percentage of carriers with a PGV in the prospective cohort.
